# Artificial Intelligence-Based Clustering and Characterization of Parkinson’s Disease Trajectories

**DOI:** 10.1101/2022.08.15.22278776

**Authors:** Colin Birkenbihl, Ashar Ahmad, Nathalie J Massat, Tamara Raschka, Andreja Avbersek, Patrick Downey, Martin Armstrong, Holger Fröhlich

## Abstract

Parkinson’s disease (PD) is a highly heterogeneous disease both with respect to arising symptoms and its progression over time. This hampers the design of disease modifying trials for PD as treatments which would potentially show efficacy in specific patient subgroups could be considered ineffective in a heterogeneous trial cohort. Establishing clusters of PD patients based on their progression patterns could help to entangle the exhibited heterogeneity, illuminate clinical differences among patient subgroups, and identify the biological pathways and molecular players which underlie the evident differences. Further, stratification of patients into clusters with distinct progression patterns could help to recruit more homogeneous trial cohorts. In the present work, we applied an artificial intelligence-based algorithm to model and cluster longitudinal PD progression trajectories from the Parkinson’s Progression Markers Initiative. Using a combination of six clinical outcome scores covering both motor and non-motor symptoms, we were able to identify specific clusters of PD that showed significantly different patterns of PD progression. The inclusion of genetic variants and biomarker data allowed us to associate the established progression clusters with distinct biological mechanisms, such as perturbations in vesicle transport or neuroprotection. Furthermore, we found that patients of identified progression clusters showed significant differences in their responsiveness to symptomatic treatment. Taken together, our work contributes to a better understanding of the heterogeneity encountered when examining and treating patients with PD, and points towards potential biological pathways and genes that could underlie those differences.

## Introduction

Parkinson’s disease (PD) is an age-associated neurodegenerative disorder that affects approximately seven million people worldwide. Alongside the cardinal motor symptoms of bradykinesia, rigidity, tremor, and postural instability [1], PD patients suffer from a wide range of non-motor symptoms such as sleep disturbances, psychosis, cognitive impairment, and mood disorders [2]. Currently there are no disease modifying treatments available for PD and present medications (e.g., L-DOPA) only offer limited symptomatic benefits. Designing and conducting clinical trials to test putative disease-modifying treatments is complicated due to the high inter-individual variability of disease progression rates [3-5]. Therefore, understanding the different biological mechanisms that drive differential disease progression is vital to ultimately pave the way for personalised therapies and can help to identify novel target candidates for therapeutic intervention.

Previous attempts to identify PD subtypes focused on ad-hoc classification of the motor characteristics of tremor (tremor dominant sub-type) and postural instability (postural instability and gait dominant sub-type) [1]. Similarly, age at disease diagnosis has been used to classify PD patients into Late Onset PD and Young Onset PD [3]. However, given the broad and complex range of PD symptoms, single-variable subtyping approaches are unlikely to capture the complexity of patients’ progression. Here, data-driven multivariate approaches using, for example, cluster analysis [5] offer a promising opportunity to overcome these limitations.

The foundation for such multivariate subtyping approaches is built through multi-modal longitudinal data provided by observational cohort studies such as the Parkinson’s Progression Markers Initiative (PPMI) [6]. PPMI data has been previously used to identify patient subtypes based on cross-sectional imaging data and cerebrospinal fluid biomarkers at study baseline [2,7]. Only a few studies have focused on disease progression which requires the use of longitudinal follow-up data. This aspect was partially addressed by Faghri *et al*. [8] using PPMI data at 48 months follow-up. The authors identified three PD subtypes using non-negative matrix factorisation. Still, their approach was unable to discern these subtypes with respect to the slope of progression. In this context, recently published neural network-based approaches make it possible to cluster entire longitudinal patient trajectories [9,10]. However, these studies did not explore the biological underpinning of the subtypes nor did they consider how their patients differed in their clinical appearance or their response to treatment.

In this work, we identified three clusters of PD progression in a purely data-driven manner based on multivariate longitudinal trajectories of *de novo* PD patients from PPMI. In doing so, we considered repeated measurements of a combination of several motor and non-motor scores over time. Furthermore, we used machine learning to identify biological pathways, genetic variations, and key clinical symptoms associated with the respective clusters. Finally, we investigated potential differences in the loss of dopaminergic neurons across clusters and further revealed cluster-specific responses to symptomatic L-DOPA treatment. Our findings contribute to a deeper understanding and characterisation of the heterogeneous mechanisms at play within PD and offer the opportunity to define novel drug targets.

## Results

### Multivariate time series analysis identifies three patient clusters with distinct progression profiles

By clustering the time series data of 407 *de novo* PD patients from PPMI (267 male, 140 female) using our previously published artificial intelligence-based VaDER approach [10], we identified three groups of PD patients with distinct progression profiles (**Supplementary Section S1, Fig. S1**). The clustering was conducted based on the multivariate progression of six key clinical assessments of PD symptoms over the course of up to 60 months: the UPDRS 1, 2, and 3 (off treatment) [11], tremor dominant score (TD), postural instability and gait disorder score (PIGD), and the Epworth sleepiness scale (ESS).

The three resulting clusters contained ‘moderate’-progressors (n = 230), ‘fast’-progressors (n = 53), and ‘slow’-progressors (n = 124), respectively. **Table 1** provides summary statistics of patients from each cluster at study baseline. We found significant differences between the average age at study baseline of the slow progressors and the two other respective subtypes (t-test ‘slow’ versus ‘fast’, p<0.013; ‘slow’ versus ‘moderate’, p<0.019; ‘moderate’ versus ‘fast’, p>0.32). In contrast, no significant difference was observed in the elapsed time from initial diagnosis to study baseline (pairwise U-tests between all three subtypes, p>0.3), or distribution of Hoehn and Yahr stages (*χ*^2^-test, p>0.15). Furthermore, we detected no significant differences in the distribution of biological sex (*χ* ^2^-test, p>0.15) and the start of symptomatic therapy (**Fig. S2**).

**Table 1:**
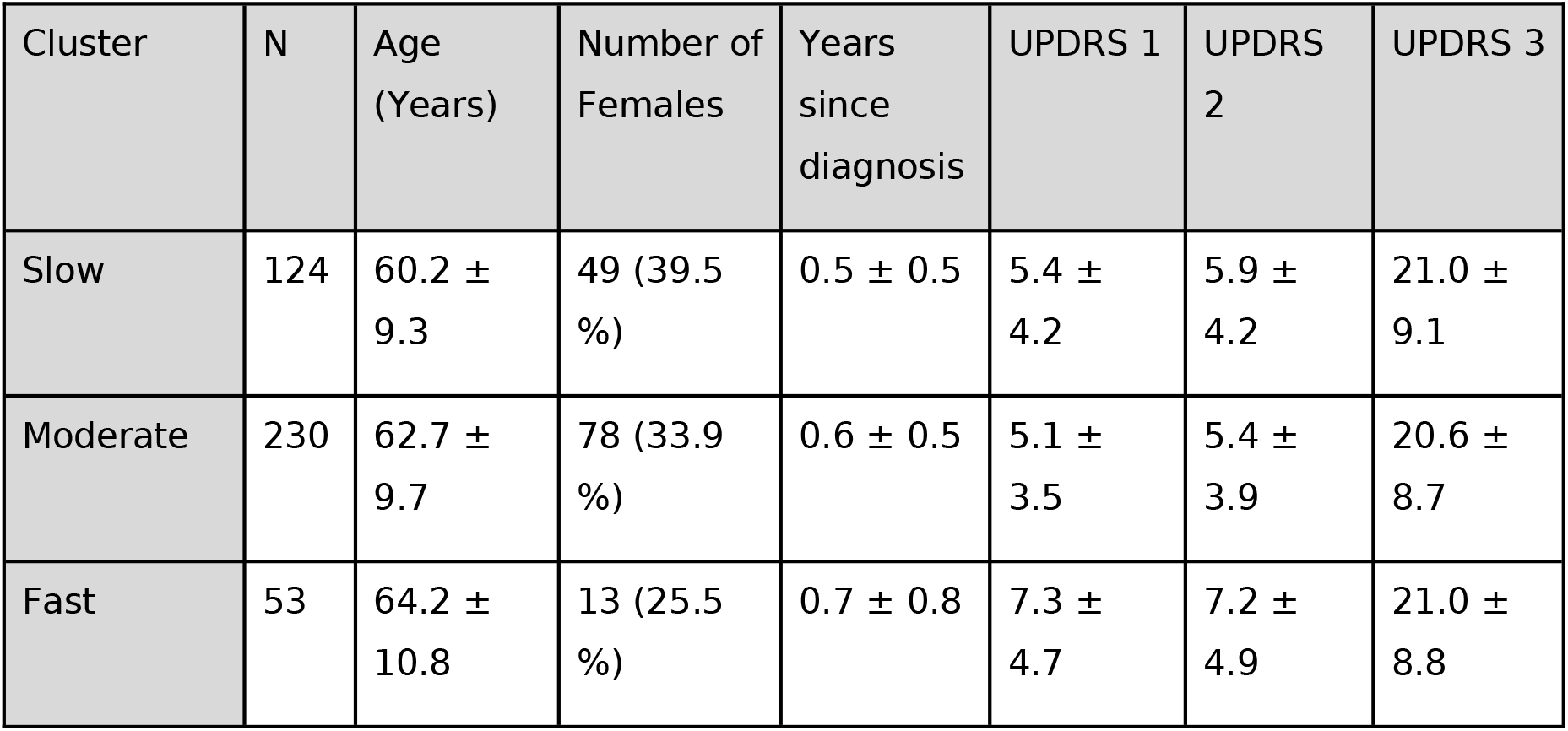
Summary statistics of patients per subtype at study baseline. Presented is the mean and standard deviation of variables as well as the percentage of females per subtype. N, Number of patients per subtype.

The mean univariate progression trajectories of these clusters along with their 95% confidence intervals are depicted in **Fig. 1**. Although the clustering was conducted on multiple outcome measures, we observed a clear separation of clusters across all selected variables except for the TD score between ‘fast’ and ‘moderate’ progressors. While ‘fast’ and ‘moderately’ progressing subtypes displayed a clear increase of symptoms over the covered 60 month interval already starting from baseline and month 12 respectively, ‘slow’-progressors experienced almost no significant symptom worsening across scores until month 24.

**Figure 1:**
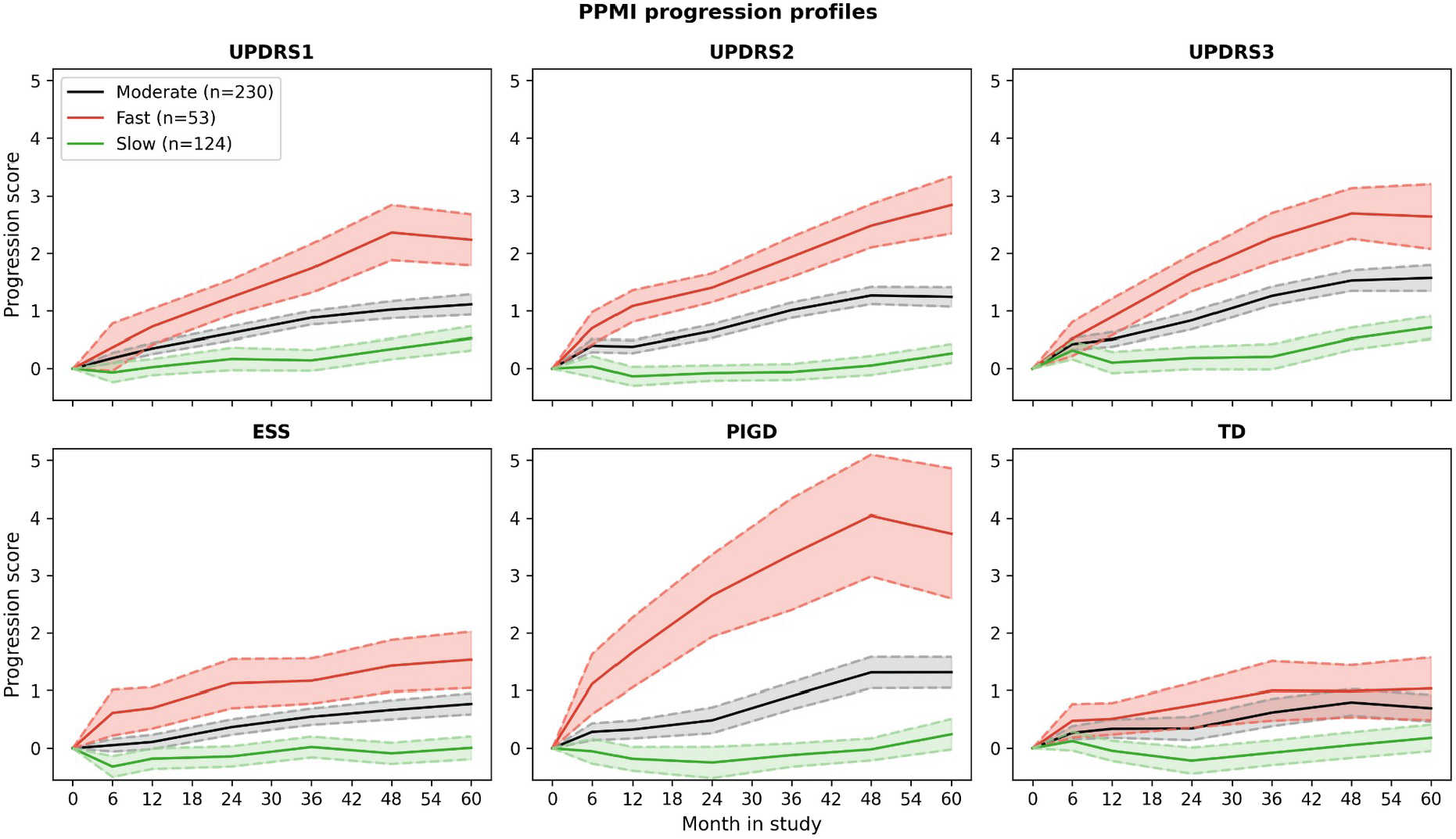
Mean trajectories of the three different progression clusters. Dashed lines depict the 95% confidence interval of the respective trajectory. Confidence intervals grow larger with time as more patients drop-out of the study. The progression score depicted on the y-axis represents the relative change to study baseline normalised by the standard deviation of the respective variable. UPDRS refers to the UPDRS testing battery, ESS to the Epworth Sleepiness Scale, PIGD to the Postural Instability Gait Disorder, and TD to the Tremor Dominant Score.

### Characterisation of PD clusters suggests longitudinal differences in dopaminergic deficiency

The differences in motor symptom progression rates across subtypes **(Fig. 1)** were mirrored by significant differences in the age-adjusted trajectories of DaTSCAN measurements, which were available until month 48: the rates in loss of specific-binding ratio (SBR) signal were greater for both the ‘fast’ and ‘moderate’ progressing subtypes in the caudate region when compared to the ‘slow’ progressing group (signal loss of -0.0033 SBR unit/month, 95% CI [-0.0055, -0.0011], p = 0.004 for the ‘fast’ group, and of -0.0019 SBR unit/month, 95% CI [-0.0032, -0.0003], p = 0.01 for the ‘moderate’ group). No significant difference at the 5% level was observed between the ‘fast’ and ‘moderate’ progressing groups **(details in Supplementary Section S3)**. The difference in rate of dopaminergic loss between the ‘fast’ and the ‘slow’ progressing clusters was seen equally in the ipsilateral (signal loss of -0.0034 SBR unit/month, 95% CI [-0.0056, - 0.0008], p = 0.008) and the contralateral (signal loss of -0.0032 SBR unit/month, 95% CI [-0.0057, -0.0008], p = 0.007) sides of the caudate region. In contrast, the difference in rate of progression between the ‘moderate’ and the ‘slow’ progressing subtypes was stronger in the contralateral side (signal loss of -0.0022 SBR unit/month, 95% CI [-0.0038, -0.0006], p = 0.006) as compared to the ipsilateral (signal loss of -0.0022 SBR unit/month, 95% CI [-0.0030, +0.0002], p = 0.07) sides of the caudate region. No significant difference in SBR rates were observed in the putamen, and changes in the striatum were intermediary between those observed in the caudate and the putamen.

### Machine learning revealed associations between clusters and underlying biology

To discover further associations between the identified progression clusters and clinical as well as biomarker and genetic variables, we developed machine learning models based on patients’ baseline visit data. Additionally, we built a second version of these models that further included 3 months follow-up data, both in the form of raw values and variables’ change relative to baseline. The variables included into the models comprised demographic and clinical data (86 variables at baseline; 217 including 3 month follow-up), CSF biomarkers (amyloid beta, phosphorylated tau, total tau), blood serum transcriptomic data (7 variables), 3472 SNPs gained through a linkage disequilibrium analysis of an initial set of 145 PD associated SNPs obtained from DisGeNET [12], and brain region specific DaTSCAN (5 variables). We also calculated burden-scores for biological pathways stemming from Kegg [13], Reactome [14], and NeuroMMSig [15] (36, 10, and 12 pathways, respectively). These scores were based on the SNP data of each respective patient and described the amount of genetic variation affecting a pathway (see Method section for details). A full list of all variables is presented in the **Supplementary Spreadsheet**.

The machine learning algorithm of choice was a sparse gradient LASSO [16]. We developed three distinct models, each discriminating one of the clusters from the respective other two (i.e., one versus rest approach). The significance of the most strongly associated variables was then determined by bootstrapping each model 200 times and investigating whether the resulting confidence intervals (CI) of standardised coefficients contained zero. CIs were Bonferroni-corrected to account for multiple testing. Further methodological details are described in **Supplementary Section S4**.

The built models revealed several significant associations between measured variables and progression clusters, which were interpretable from a clinical as well as a biological point of view.

### Progression clusters are associated with distinct symptoms and genetic loci

The coefficients of each machine learning model highlight how specific variables influence the probability that a patient belongs to a particular cluster. For interpretability, we focused on significant positive interactions (i.e., variables that increase the chance of belonging to the respective cluster; **Fig. 2A-C**).

**Figure 2:**
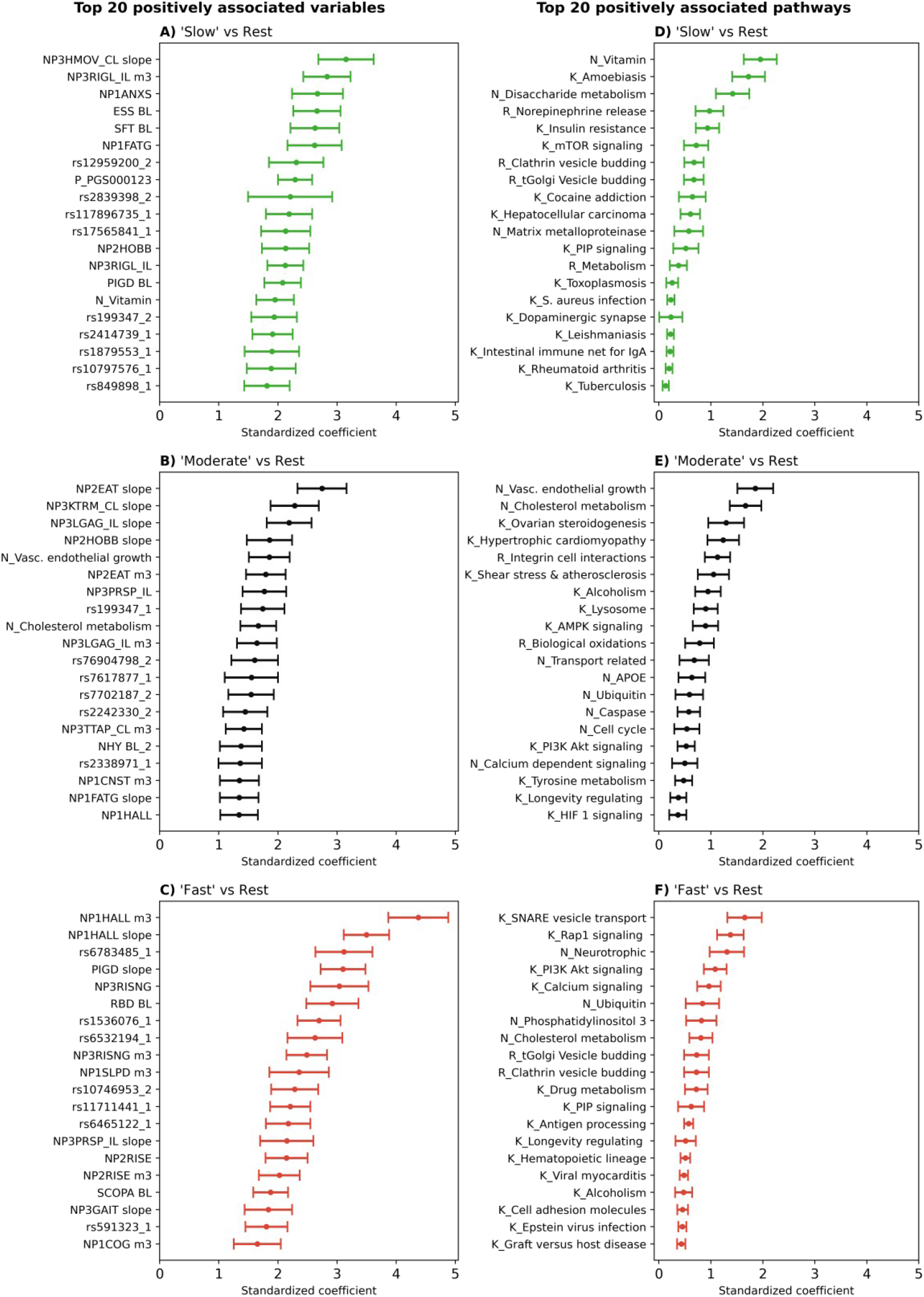
Top 20 variables associated with the respective progression cluster (sparse group LASSO using baseline data + 3-month follow-up). The plots show the standardised coefficient together with their Bonferroni-corrected 95% confidence intervals for each variable. A stronger positive coefficient value in the plot indicates a higher likelihood of a patient to belong to the respective cluster. A corresponding plot for baseline data only is shown in **Fig. S7. A-C**, most associated variables for ‘slow’, ‘moderate’ and ‘fast’ progression. Variables ending in ‘slope’ indicate the slope of the corresponding score measured 3 months after baseline. The number after SNP IDs indicates the number of non-reference alleles. **D-F**, most associated biological pathways. Pathways starting with ‘K_’, ‘R_’, or ‘N_’ originate from Kegg, Reactome, and NeuroMMSig, respectively. PGS denotes polygenic risk scores.

The variable most strongly associated with ‘fast’ PD progression was the presence and severity of hallucinations at the 3 month follow-up visit (NP1HALL m3, 95%CI [3.91, 5.0]), with the increase in experienced hallucinations following in third position (NP1HALL slope, 95%CI [3.07, 3.9]). In fourth position, the increase in postural instability and gait disorder severity over the first 3 months was found (PIGD slope, 95% CI [2.73, 3.55]). Additionally, ‘fast’ progressing patients experienced more difficulties when rising from a lying or sitting position compared to the other two subtypes (95% CI: NP3RISNG [2.56, 3.63], NP3RISNG m3 [2.16, 2.98], NP2RISE m3 [1.9, 2.65], NP2RISE [1.8, 2.64]). REM sleep behaviour disorder (RBD) proved to be another association for ‘fast’ progression (95% CI [2.33, 3.24]). Furthermore, several SNPs (rs6783485 - LOC105377110, rs1536076 - SH3GL2, rs6532194 – chromosome 4:89859751, rs11711441 - chromosome 3:183103487, and rs591323 - LOC105379297) were found to be among the top 20 associated variables for ‘fast’ progression. Notably, all these SNPs were taken from DisGeNET, because of their known association to PD according to GWAS studies. In all cases, the non-reference-allele increased the risk of ‘faster’ PD progression.

‘Slow’ PD progression was associated with increasing difficulties when performing the hand movement task of the UPDRS (NP3HMOV slope 95% CI [2.93, 3.38]). Furthermore, a series of highly associated variables were connected to daytime sleepiness (ESS 95% CI [2.27, 3.06]) and general fatigue (NP1FATG 95% CI [2.16, 2.97]). Patients of the ‘slow’ cluster also suffered more often from anxiety (95% CI: NP1ANXS [2.15, 2.93]; NP1ANXS m3 [0.89, 1.53]) and were the only subtype which showed a significant positive association with depression, albeit the coefficient remained rather small (geriatric depression scale 95% CI [0.1, 0.65]). Additionally, better semantic fluency was also connected to ‘slower’ disease progression (SFT 95% CI [2.06, 2.84]). With regard to motor symptoms, ‘slow’ progression was associated with rigidity of the ipsilateral extremities at baseline, month 3, and their relative increase in severity (95% CI: NP3RIGL_IL m3 [2.23, 3.09]; NP3RIGL_IL [1.74, 2.54]; NP3RIGU_IL [1.0, 1.61]). Further, we found a significant positive association of the polygenic risk score PGS000123 [17] and multiple genetic loci with the probability to belong to the ‘slow’-progressors. SNPs rs17565841 (OCA2), and rs12959200 (chromosome 18:73599819) placed among the top 10 associations (95% CI: [2.11, 2.71], [1.95, 3.05], [1.91, 2.77], respectively). Once again, these SNPs were taken from DisGeNET because of their known association to PD according to GWAS studies

For ‘moderate’ disease progression, the strongest association was the worsening of performing the eating task of the UPDRS over the first 3 month (NP2EAT slope 95% CI [2.3, 3.08]). Further, reduced agility in the ipsilateral leg was associated with ‘moderate’ progression (95% CI: NP3LGAG_IL slope [1.79, 2.55]; NP3LGAG_IL m3 [1.36, 2.06]). With rs76904798 (chromosome 12:40220632), rs199347 (GPNMB), rs7702187 (SEMA5A), and rs7617877 (LINC00693), we identified several PD associated SNPs which raised the probability for patients to belong to the ‘moderate’ subtype.

A comprehensive view on all variables and their coefficients can be found in the **Supplementary Spreadsheet**.

### Genetic burden scores connect the heterogeneity in PD progression to biological pathways

Several biological pathways and genes could be associated with the respective clusters **(Fig. 2 D-F)**. The ‘fast’ cluster was highly associated with higher genetic burden in the Kegg ‘SNARE vesicle transport’ pathway (95% CI [1.25, 1.92]), the ‘Rap1 signalling’ pathway (95% CI [1.1, 1.71]), and NeuroMMSig’s ‘neurotrophic’ subgraph (95% CI [1.25, 1.92]). The patients of the ‘moderate’ cluster were linked to the ‘cholesterol metabolism’ subgraph (95% CI [1.56, 2.25]) and ‘vascular endothelial growth factor’ subgraph (95% CI [1.42, 2.12]) originating from NeuroMMSig. The ‘vitamin’ and ‘disaccharide metabolism’ subgraphs from NeuroMMSig, and Kegg’s ‘amoebiasis pathway’ were discovered as strongly associated with the ‘slow’ progressing clusters (95% CI: [1.6, 2.22], [1.04, 1.66], and [1.14, 1.86], respectively). A list of all mappings between pathways, genes and SNPs can be found in the **Supplementary Spreadsheet**.

### Identified clusters show differences in response to motor symptom therapy

After observing that potentially different biological pathways were involved in the PD pathology of each cluster, we investigated whether the clusters also differed in their response to symptomatic treatment for motor symptoms. To this aim, we selected participants who had initiated Levodopa or Dopamine agonist symptomatic treatment between month 6 and month 9 after baseline and assessed whether progression as measured by UPDRS 3 sub-score differed by PD cluster. We separately analysed the ‘ON’-state UPDRS 3 score data, in which patients are examined approximately one hour after taking medication (**Fig. 3**), and the ‘OFF’-state UPDRS 3 score data (**Fig. S11**), in which patients were examined at least 6 hours after the last dose, which is considered sufficient time for washing off the effect of levodopa or dopamine agonists [18]. Methodological details and statistical plots can be found in **Supplementary Section S6**.

**Figure 3:**
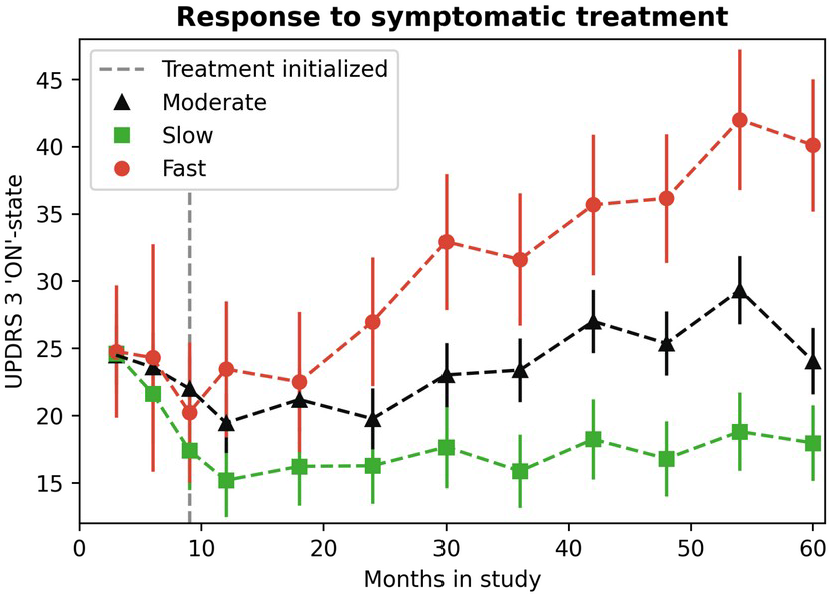
Differential response to symptomatic treatment. Effect plot of modelled UPDRS 3 ‘ON’-state score progression prior to and after the initiation of Levodopa or Dopamine agonist in patients who initiated therapy between 6 and 9 months post-baseline using a longitudinal LMEM with time fitted as a categorical variable and baseline score fitted as a covariate. The error bars represent the 95% confidence intervals, based on standard errors computed from the covariance matrix of the fitted regression coefficients.

Although initially all three PD clusters responded similarly to symptomatic treatment by stabilising their motor scores in the first 9 months after treatment initiation (i.e. 9 - 18 months post-baseline, **Fig. 3, Fig. S11**), we observed that patients in the ‘red’ cluster continued to progress fastest and all three clusters had significantly different UPDRS 3 scores in ‘ON’ and ‘OFF’-states at 30 months after baseline (i.e. 21 months post-symptomatic treatment initiation) from each others, i.e. the 95% CIs did not overlap. At the same time we could neither observe any significant differences in type of treatment nor Levodopa equivalent daily dose across clusters (**Tables S1, S2**).

## Discussion

In this work, we identified three distinct PD progression clusters dividing patients into ‘slow’, ‘moderate’, and ‘fast’-progressors. This clustering built on the multivariate trajectory of six clinical variables rather than a single univariate outcome. Evaluating potential confounders that could have biased the clustering showed no significant differences of biological sex, disease duration, and Hoehn & Yahr stages across clusters. A machine learning model further identified significant associations between clinical measurements taken at study baseline (optionally including 3 months follow-up data), genetic features, biological pathways, and the different progression clusters of patients. Several distinct SNPs and biological mechanisms could be associated with each cluster. Analysis of the observed associations provides insights into the heterogeneity of PD progression and the distinct biological pathways promoting it. Further analysis revealed that patients in different clusters responded differently to symptomatic treatment and displayed significant differences in dopaminergic cell loss. Altogether this makes it improbable that our clustering is just a consequence of patients being in different disease stages at study baseline.

### Interpretation of significant associations between variables and PD progression clusters

Our machine learning models identified that measurements taken early in the disease course already show significant associations with the longitudinal progression of PD’s motor and non-motor symptoms.

In the ‘fast’-progressing clusters, the presence of psychotic symptoms in the form of hallucinations or delusions was found as the strongest association. Indeed, hallucinations can already be observed in newly diagnosed patients [19] and experiencing such visual or auditory hallucinations was established to be one of the most notable risk factors for increased mortality [20] and earlier placement in care homes [21]. These findings could, on the one hand, be explained by the difficulties of living with psychosis but, on the other, also point towards a faster disease progression in general. In this context, the association between RBD and our ‘fast’ progressing cluster is noteworthy, as RBD is one of the major risk factors for hallucinations [22] and was also hypothesised to be an early sign of faster disease progression [23]. Furthermore, RBD has been connected to reduced striatal dopaminergic activity [24], which is in line with our observations for the ‘fast’ progressing cluster. In concordance, Wang et al. discovered slower and faster progressing subtypes based on brain pathology with the faster subtype showing increased RBD and decreased dopaminergic brain efficiency in the caudate and putamen at study baseline [25]. In another subtyping effort by Fereshtehnejad et al. a ‘diffuse malignant’ PD cluster was described that showed faster disease progression and was characterised by lower CSF amyloid beta values [26]. Indeed, our ‘fast’ progressing cluster was also associated with lower amyloid beta in CSF, however, considerably older and more affected by hallucinations than the presented ‘diffuse malignant’ subtype. Since the investigated PPMI patients were de novo PD patients, the significant difference in age across clusters at baseline added further evidence to a previously discovered trend that patients with later disease onset often experience faster progression [27,28].

The ‘slow’ cluster showed strong associations with non-motor symptoms such as fatigue, sleepiness, and anxiety. While these symptoms have received increasing recognition in recent years, they remain poorly understood aspects of PD [2] and little is known about disease progression in patients that suffer from them.

Previous case series reported on several associations between slower disease progression and attributes we found to be significant associations with what we called ‘moderate’ progression [29]. Here, it was described that patients with predominantly worsening tremors, younger age, and no indication of PGID showed reduced disease progression. Related to that, our results further underline the notion that postural instability during the first three years post diagnosis represents an untypical feature for PD [30]. In both more extremely progressing clusters (ie., ‘slow’ and ‘fast’), PIGD related variables showed significant positive association. However, the majority of patients belonged to the cluster displaying ‘moderate’ progression where no positive PIGD association was discovered.

Only slight differences in global cognitive performance as measured by the Montreal Cognitive Assessment (MoCA) could be found among the clusters. This could be due to the comparably early time point of assessment (approximately one month after PD diagnosis for most patients), since only subtle cognitive changes are observable in the PPMI cohort over the first 5 years [2]. However, semantic fluency was among the strongest associated variables with ‘slow’ progression, indicating that this cluster could be more stable with respect to cognitive performance.

### PD progression clusters are associated with distinct biological pathways and gene mutation load

With the inclusion of available genetic data into the models, we were able to identify distinct biological pathways that were associated with the different clusters. This opens up the opportunity of not only identifying new therapeutic targets, but targets that may be positioned more effectively within certain subgroups of patients.

The pathway most predominantly associated with ‘fast’-progression was the Kegg ‘SNARE interactions in vesicular transport’ pathway. Vesicle dysfunction is a known phenomenon in the pathogenesis of PD, the targeting of related proteins (including SCNA and LRRK2) has been discussed for several years now [31] and there are multiple lines of supporting evidence for the role of this pathway in PD. In this pathway, the retrieved SNPs predominantly mapped to genes encoding for vesicle associated membrane proteins (VAMP2, VAMP4) and syntaxins (SXT4, and SXT1B). VAMP2 interacts with SXT1 in the neuronal synapse and is important for vesicle fusion and neurotransmitter translocation [32,33]. VAMP4 and syntaxins interact with LRRK2 [34], a major PD risk factor and potential drug target in which mutations promote a PD phenotype [35], with respect to retrograde and post-Golgi signalling. Both VAMP2 and SXT1 showed diagnostic potential in blood-based biomarker studies for PD [36].

The second strongest association found for fast progressors was the ‘Rap1 signaling’ pathway which is involved in the nigrostriatal dopaminergic pathway in medium spiny neurons [37]. Again, multiple lines of supporting evidence lend biological support to the role of this pathway, including the position of the vascular endothelial growth factor (VEGFA) gene in the pathway, that has been shown to protect dopaminergic neurons from cell death. VEGFA has been discussed as a potential target for treating PD [38] and a recent study suggests blocking of VEGFA to prevent blood-brain-barrier disruption, which has been implicated in several neurodegenerative diseases, including PD [39].

Furthermore, this pathway involves several fibroblast growth factors (FGF5, 10, and 20), with FGF20 also being a prominent entity in the ‘Neurotrophin’ mechanism listed in NeuroMMSig (the third most associated pathway for ‘fast’ progression). The FGF gene family has also been associated with neuroprotection and neurogenesis, partially by triggering PI3K-AKT signalling which also occurred among our highly associated pathways with respect to ‘fast’ PD progression [40].

Taken together, it can be postulated that severe perturbations in Golgi vesicle transport that eventually cause apoptosis, in combination with a reduced neuroprotection and neurogenesis to replace damaged cells might promote a ‘fast’ progressing form of PD.

The ‘moderately’ progressing cluster was mainly associated with NeuroMMSig’s ‘Vascular endothelial growth factor’ and ‘Cholesterol metabolism’ pathways. The former was largely defined by VEGFA which was discussed above and might indicate a common mechanism between ‘fast’ and ‘moderate’ progressors. The squalene synthase (FDFT1) was the major gene in the ‘cholesterol metabolism’ pathway to which we could map SNPs. Squalene is an antioxidant and precursor of cholesterol which is essential for synaptic functioning and has been linked to PD and α-synuclein aggregation [41]. This, along with additional supporting evidence for this pathway [42-47], could indicate that oxidative stress might play a more pronounced role in ‘moderately’ progressing PD compared to the other two subtypes.

The strongest associated pathway for the ‘slow’ progressing cluster was the ‘Vitamin subgraph’ which evolved around the solute carrier family 41 member 1 (SLC41A1). This gene is part of the PD related PARK16 locus and is associated with magnesium efflux and homeostasis which is believed to contribute to PD [48]. Furthermore, the ‘amoebiasis’ pathway was identified as the second highest associated and the connection of the underlying genes to PD has been observed previously [49]. Interestingly, we also found an association of ‘slow’ progressing PD to the ‘disaccharide metabolism’ pathway, in which GBA was a key agent. Whilst it should be noted that GBA mutation carriers were excluded from the PPMI dataset used in this analysis, three SNPs in our analysis could still be mapped to GBA, (rs2230288, rs12752133, and rs76763715) and all have been associated to an increased risk of PD [50].

### Differential response to symptomatic motor treatment across progression clusters

When the progression of motor symptoms was compared between the clusters after the initiation of Levodopa and/or Dopamine agonists, a substantial difference in the response to the symptomatic treatment was observed, which could not be explained either by medication dosage or type of therapy. Together with the observed genetic differences between clusters, our results strongly suggest that the identified progression clusters represent an inherent property of the disease. Notably, differential response to symptomatic treatment for PPMI de-novo PD cohort participants with fastest motor progression was also reported in [51], and by Lawton *et al*. using data from the Tracking Parkinson and Oxford Parkinson’s Disease Centre Discovery cohort [52].

### Limitations

When interpreting the genetic data, it should be noted that our SNP inclusion was hypothesis driven based on prior evidence for an association with PD. Nevertheless, the work presented highlights the ability of the models to discriminate between molecular pathways involved in the different clusters, and the importance of genetic data in PD. The availability of larger datasets with attached genome wide genetic data would support a more hypothesis generating approach and potentially uncover novel mechanisms.

It should be mentioned that PPMI as a primary data source for our analysis is an observational study, in which patients are treated according to best clinical routine practice. The treatment itself is not monitored precisely, and the actual compliance of patients to medications remains unclear.

Finally, our approach relies on a clinical diagnosis of PD. Potential misdiagnosis of patients as PD could therefore bias the results.

## Conclusion

Using our clustering approach, we show that PD patients can be divided into ‘slow’, ‘moderate’, and ‘fast’-progressors based on the relative change of symptoms over the time course of the study. These groups not only show differences in the progression rates of clinical symptoms but also differ in the rate of dopaminergic cell loss, and importantly respond differently to symptomatic treatment. An analysis of whole genome sequencing data also suggests that genetic and mechanistic differences underpin these groupings. Currently, several agents are being tested in the clinic for their ability to slow disease progression but running such trials in a group of patients containing individuals with very different progression rates is fraught with difficulty. In the PPMI cohort that we used in this work, we identified 124 of 407 patients as slow progressors, and these patients showed no worsening of any symptom for at least 24 months. Given that current disease modifying trials in PD do not exceed two years, one can expect about a third of the patients to show no symptom worsening for the duration of the trial, provided that PPMI can be regarded as a representative PD study. As disease modifying treatments do not aim to improve symptoms but to slow down their worsening then the presence of a significant number of slow progressors who will not deteriorate during the trial will make it very difficult to observe disease slowing in a mixed population even with a highly effective treatment.

Future work is needed to further validate our established PD progression clusters ideally with the help of a larger study where similar data modalities as in PPMI are measured in de-novo PD patients.

## Materials and Methods

### Dataset and patient selection criteria

We selected 407 de-novo PD patients from the PPMI dataset. Our inclusion criteria were: age older than 30 years, Hoehn and Yahr stage of 1 or 2, recent PD diagnosis, and untreated by anti-PD medication (patient in the off-state according to the PPMI data). Furthermore, we used only patients with at least 48 months of follow-up. PPMI acquired informed consent to data collection and sharing from all participating individuals and got ethical approval. Ethical guidelines on human data collection were adhered to.

### Preprocessing by calculating progression scores

To cluster patients, the selected variables were transformed into “progression scores” that capture each variable’s change relative to baseline. We calculated these progression scores by subtracting the baseline value from the value measured at each respective time point and dividing the result by the variables standard deviation at baseline. When training the machine learning models, the raw baseline (or month three) measurements were taken and standardised or one-hot-encoded (ie., in contrast to the clustering they were progression agnostic).

### Multivariate clustering of clinical trajectories

Optimal hyperparameters for the VaDER model were found following the procedure described in [10]: We evaluated several possible models using a varying set of hyperparameters (including the number of sought clusters) and, finally, selected the hyperparameters which led to the best model performance. The performance of the model was quantified by comparing the prediction strength of the model against a random subtyping of the same data. We selected the smallest number of clusters that showed a significant difference to a random clustering with respect to the achieved prediction performance **(Fig. S1)**. The clustering was repeated 20 times and the final subtypes were assigned based on a consensus clustering across the 20 repeats. **Supplementary Section S1** provides further details, including diagnostic plots.

### Characterisation of PD progression clusters

#### Analysis of dopaminergic deficiency

DaTSCAN data were analysed for differences between PD clusters over time. Data from baseline up to 48 months was considered. Participants without DaTSCAN screening data (N = 17) were excluded from the analysis, leaving data for 390 participants. The longitudinal progression profile for individual patients in each cluster is shown in **Fig. S6**. Details about the statistical analysis are presented in **Supplementary Section S3**.

#### Response to symptomatic therapy

Patients were defined as being on symptomatic treatment, if they were taking L-DOPA, or dopamine agonists, with or without other types of motor symptom therapy such as MAO-B inhibitors at a respective visit [18]. Since a relatively highest fraction of patients started treatment at 9 months of follow-up, we focused our analysis on this time point. Altogether 44 in the ‘slow’ cluster started a symptomatic treatment at 9 months, 67 in the ‘moderate’, and 16 patients in the ‘fast’ cluster. The longitudinal progression profile using loess smoothing for individual patients in each cluster is shown in **Fig. S8**. Details about the statistical analysis including diagnostic plots are presented in **Supplementary Section S6**.

#### Analysis of whole genome sequencing data

PPMI provides whole genome sequencing (WGS) data of de novo diagnosed PD patients. To reduce the extreme high dimensionality of the WGS data while taking into account the very limited sample size, we focused only on single nucleotide polymorphisms (SNPs) with putative association to PD. More specifically, we obtained an initial list of 646 PD associated SNPs obtained from GWAS Catalogue [53], PheWas [54], and DisGeNET [12]. This list was subsequently expanded via linkage disequilibrium analysis (LD, *r*^2^ >0.8) using Haploreg [55], which also provides a gene mapping based on proximity. In addition, we employed a cis-eQTL mapping via GTex [56] to associate SNPs to genes expressed in brain tissues. Altogether 14520 SNPs were mapped to 1055 genes. In a second step, the genes were further mapped onto 12 PD specific mechanisms defined in the NeuroMMSig database [15], as well as 36 KEGG [13] and 10 Reactome [14] pathways that were significantly enriched for PD associated genes. How we calculated the pathway scores based on the selected SNPs is presented in the **Supplementary Section S4**.

## Data Availability

The authors have no permission to directly share any of the patient-level data as stated by the data usage agreement with the original data owners, namingly the PPMI study. The PPMI data used in this study can however be accessed at www.ppmi-info.org after successful access application.

## Acknowledgments

This project was partially funded by the EU-wide ERAPerMed project DIGIPD (01KU2110) and the European Union’s Horizon 2020 research and innovation program under grant agreement No. 826421, “TheVirtualBrain-Cloud”.

Data used in the preparation of this article were obtained from the Parkinson’s Progression Markers Initiative (PPMI) database www.ppmi-info.org/data. PPMI - a public-private partnership – is funded by the Michael J. Fox Foundation for Parkinson’s Research and funding partners. A list of names of all the PPMI funding partners can be found at www.ppmi-info.org/about-ppmi/who-we-are/study-sponsors/.

## Author Contributions

Designed the project: MA, HF; supervised the project: MA, HF, AAv, PD; analysed the data and implemented algorithms: CB, TR, NJM, AAh; drafted the manuscript: CB, HF, MA, PD, AAv, NJM; all authors have read and approved the manuscript.

