## Supplementary Material for "Artificial Intelligence-Based Clustering and Characterization of Parkinson’s Disease Trajectories"

### Supplementary Material: AI Based Clustering and Characterization of Parkinson's Disease Trajectories

**) This work was partially performed, while the author was affiliated with UCB Biosciences GmbH

**Multivariate time series clustering** **2**

**Characterization of clusters with respect to medication free history** **4**

**Characterization of clusters with respect to dopaminergic deficiency** **4**

**Machine learning reveals associations between variables and clusters** **11**

Sparse Group LASSO Penalised Binomial Regression 11

Training Procedure 12

Calculation of Mechanism Burden and Polygenic Risk Scores 12

**Cluster - Variable Associations Using Only Baseline Variables** **14**

**Differences in Response to Motor Symptom Therapy Across Clusters** **15**

Statistical Methodology 15

**References** **20**

#### Multivariate time series clustering

The clustering was performed using the previously published VaDER approach [1]. VaDER is a neural network based clustering approach resembling a recurrent variational autoencoder. However, in contrast to standard variational autoencoders, VaDER learns a latent multivariate Gaussian mixture model, which can be used for a probabilistic cluster assignment of patients. Another feature of VaDER is the ability to implicitly impute missing values as part of the model training.

VaDER model training and internal validation against a random clustering of the patients was performed as described in the original publication by De Jong et al. [1]. In agreement to the original publication the number of clusters k was chosen as the smallest number, where the prediction strength [2] of the true model significantly exceeded that of a null model, in which cluster memberships of patients predicted by the VaDER model had been randomly permuted (**Fig. S1**). Briefly, the prediction strength resembles a 2-fold cross-validation, in which the overall dataset is split in two equal sized halfs, one for training and one for testing. The VaDER clustering is initially performed independently on the training as well as test data. For each cluster we then measure the proportion of observation pairs in the test data that are also assigned to the same cluster by the VaDER model fitted on the training data. The prediction strength is the minimum of this quantity over the k test clusters. We refer to Tibshirani and Walther for more details to [21].

Each VaDER model was trained 20 times, starting from different random initializations. After model optimization and choosing the number of clusters a final assignment of patients to clusters was performed via consensus clustering.

Within the VaDER training model hyperparameters were tuned via a grid search. The final hyperparameters for our selected model were:

- Number of hidden layers: 2
- Number of units first layer: 128
- Number of units second layer: 8
- Learning rate: 0.0001
- Batch size: 16
- Alpha: 1.0
- k: 3

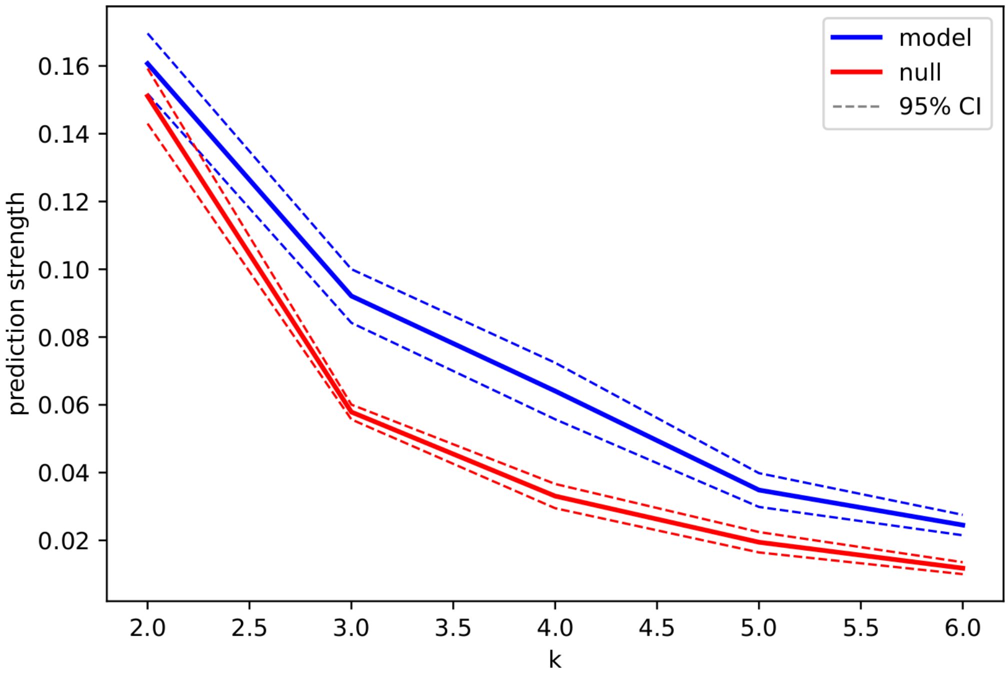

**Figure S1:** Prediction strength for the VaDER subtypes (model) compared to a random permutation of cluster memberships (null) for several numbers of clusters k. For each k 20 VaDER models starting from a random initialization were run. The plot shows the distribution of prediction strength values for each k.

#### Characterization of clusters with respect to medication free history

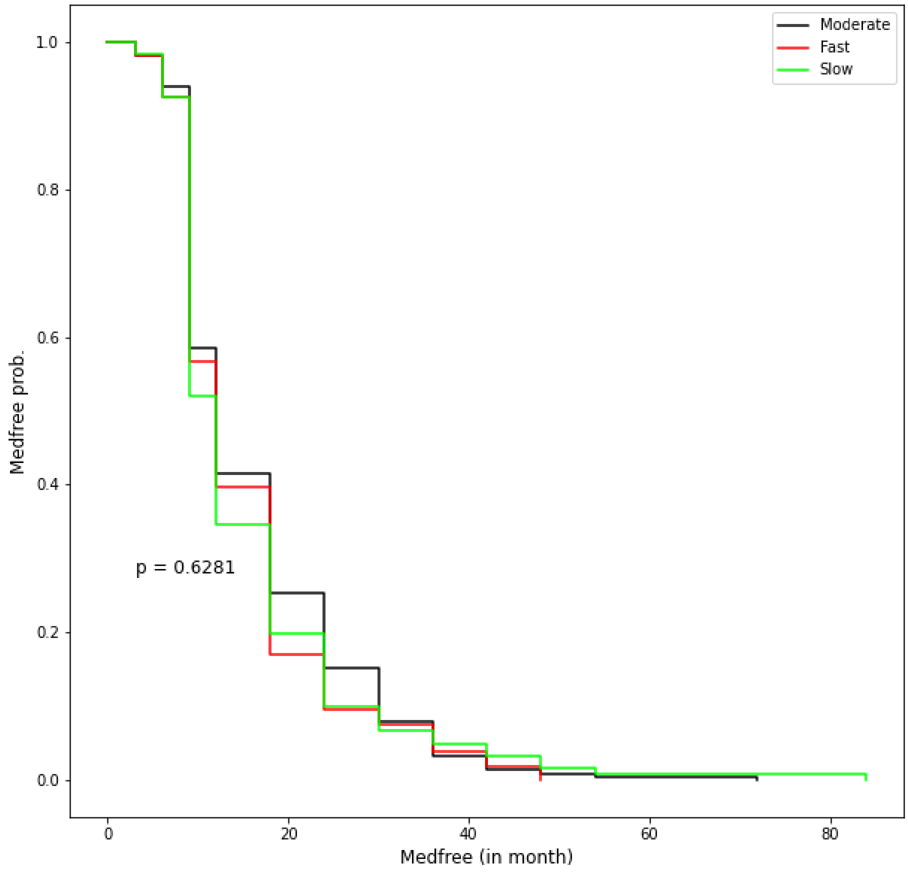


**Figure S2**: Probability of staying free of symptomatic treatment. The plot shows the time till the event "initiation of symptomatic treatment" across the different clusters. The p-value was estimated via a log-rank test.

#### Characterization of clusters with respect to dopaminergic deficiency

Loss in DaTSCAN signal was assumed (1) to be linear of the 48-month period based on the shape of individual trajectories (**Fig. S3**) and (2) not to be affected by initiation of ST intake [3]. Therefore, a single slope-LMEM was fitted to these data. Time as continuous variable, PD cluster as categorical variable and PD cluster by time as interaction were included as fixed effects into the LMEM, and a subject-specific random intercept was fitted. Baseline data was fitted as part of the dependent variable [4], [5]. The model was adjusted for age at baseline (3 categories: <55, 55-65, 65+; significant predictor of DaTSCAN signal at baseline) and for disease duration (3 categories: <0.3 year,  0.3-0.6 year, 0.6+ year; not to be a significant predictor of DaTSCAN signal at baseline).

A similar model was fitted to post-baseline data only, using baseline signal as covariate, and fitting time as a categorical fixed effect (i.e. analysis of covariance, **Fig. S4 - S6**). No imputation was performed as data were analysed using a LMEM for longitudinal data, with embedded missing at random (MAR) assumption and implicit imputation.

Mean estimates were obtained for each model parameter and corresponding 95% confidence intervals (CIs) were computed using boostrapping, and the rate of progression (slope) for each PD cluster plotted for each period against time. Reported p-values are for Satterthwaite based F-tests for difference from 0 (for baseline covariates) or for mean difference from reference category (for intercept and slopes). Diagnostic plots are shown for the model fitting time as a continuous variable (**Fig. S4)**. LMEMs were fitted separately to the caudate (whole, ipsilateral, and contralateral regions), to the putamen (whole, ipsilateral, and contralateral regions) and to the whole striatum specific-binding ratio (SBR) data.

A. Caudate

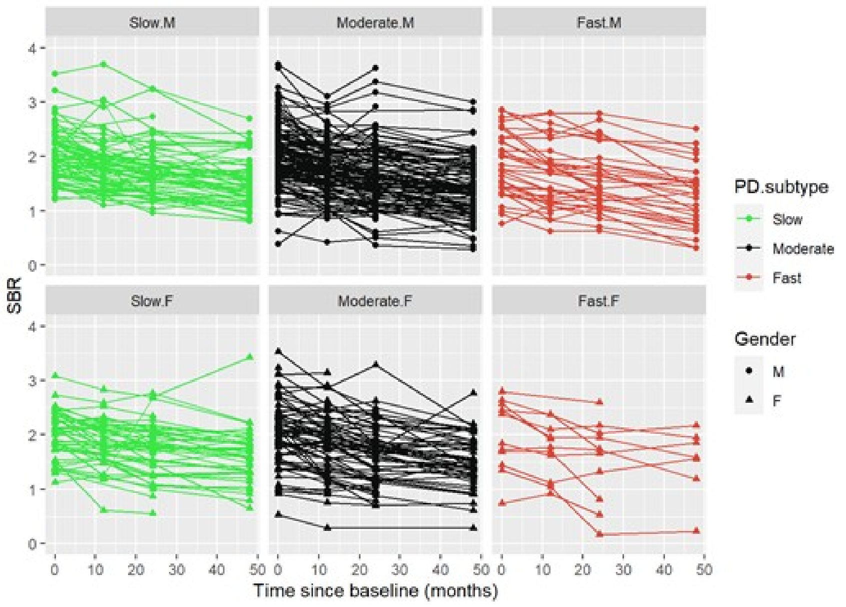


B. Putamen

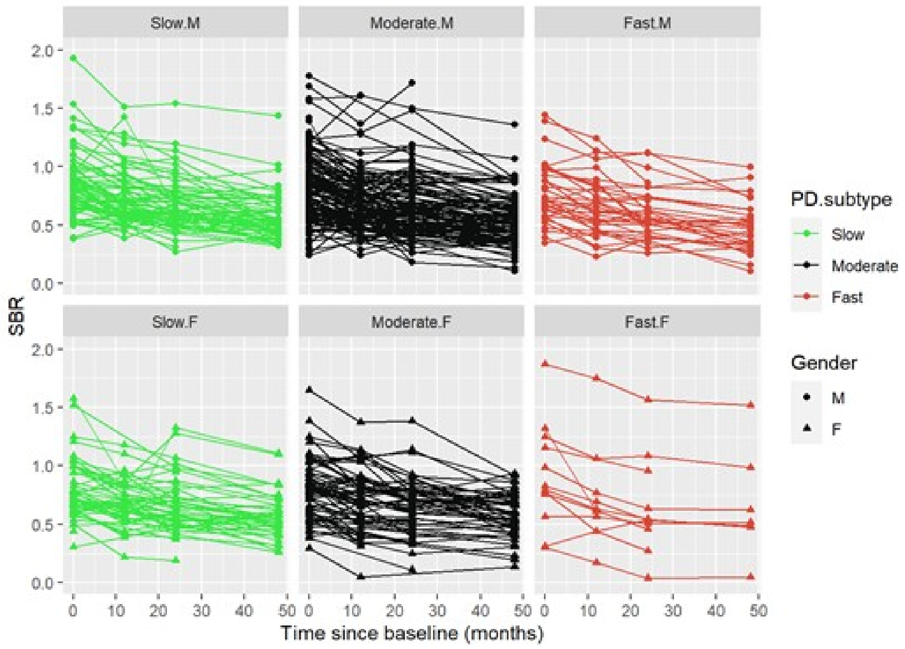


C. Whole Striatum

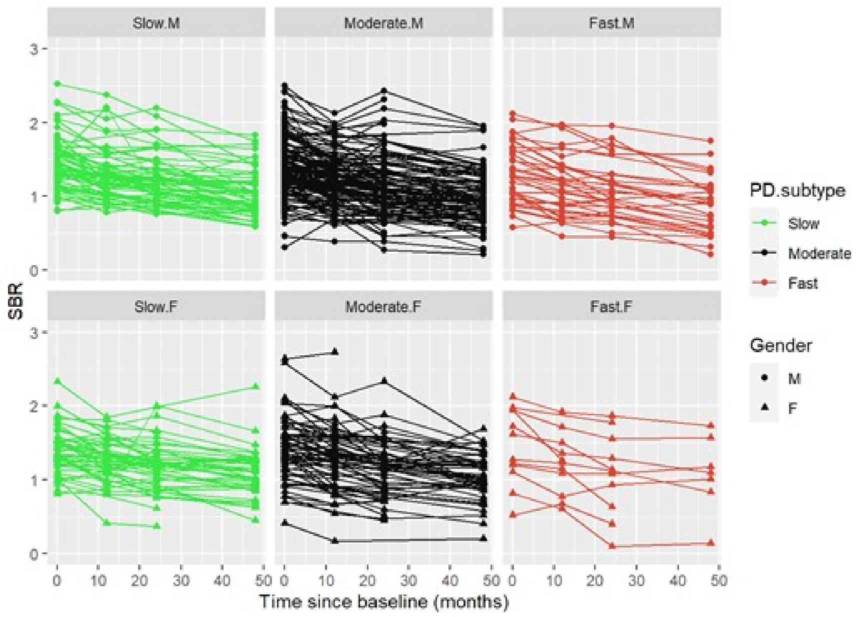


**Figure S3:** Individual line plots of DaTScan SBR signal in the caudate (A), putamen (B) and whole striatum (C) presented by gender and PD subtypes. M: Male, F: Female.

A. Caudate

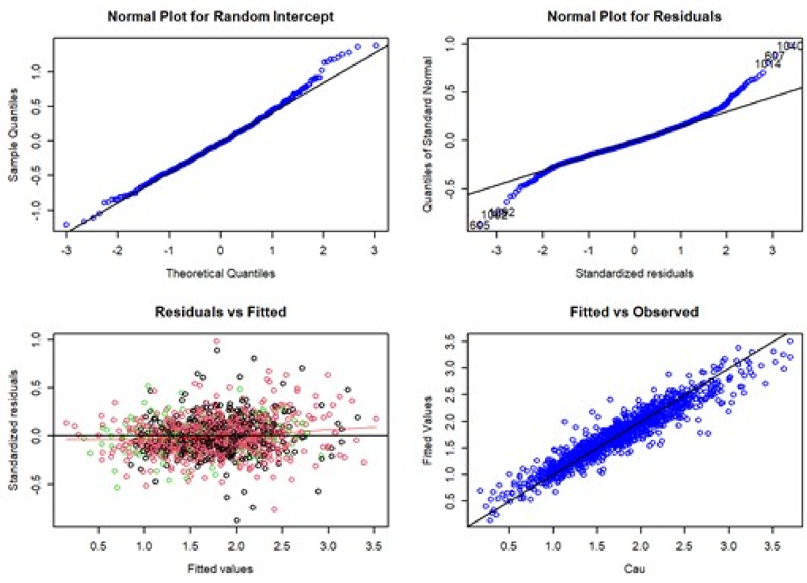


B. Putamen

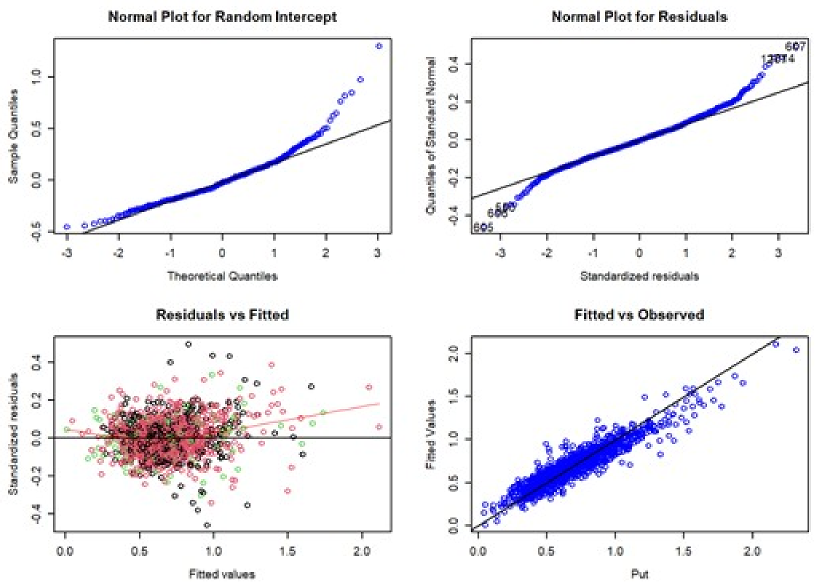


C. Whole Striatum

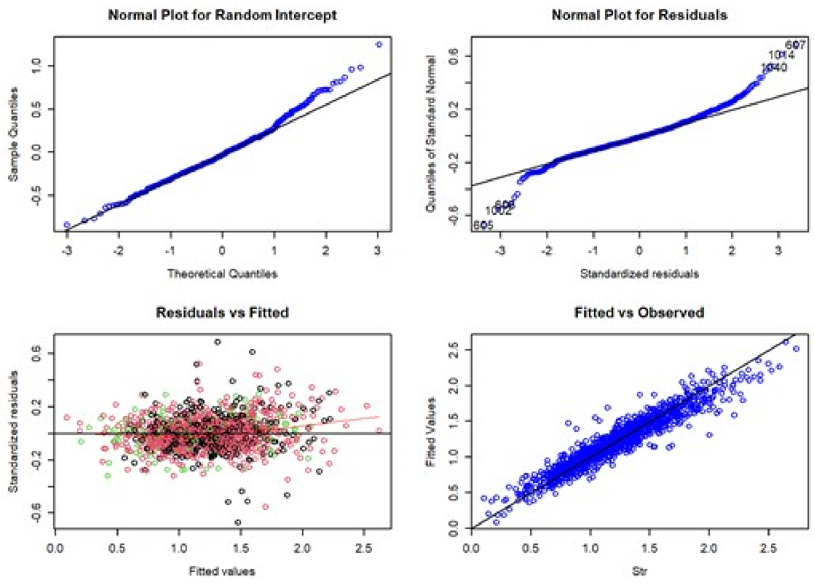


**Figure S4:** Diagnostic plots for LMEM applied to caudate (A), putamen (B) and whole striatum (C) DaTScan SBR signal where time was fitted as a continuous variable and baseline signal as response. For the ‘Residuals vs Fitted’ plot, datapoints are coloured according to the progressing group: ‘slow’ (green), ‘moderate’ (Black), and ‘fast’ (red).

A. Caudate

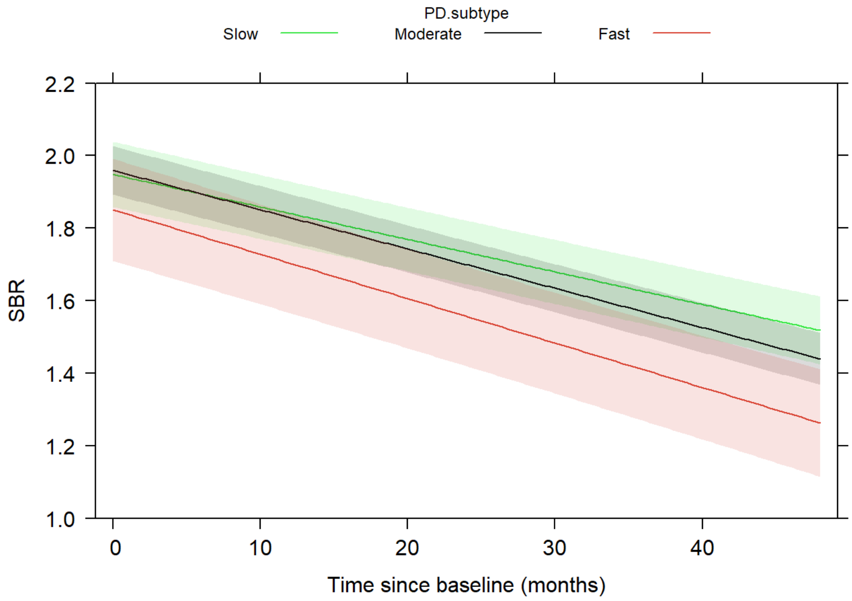


B. Putamen

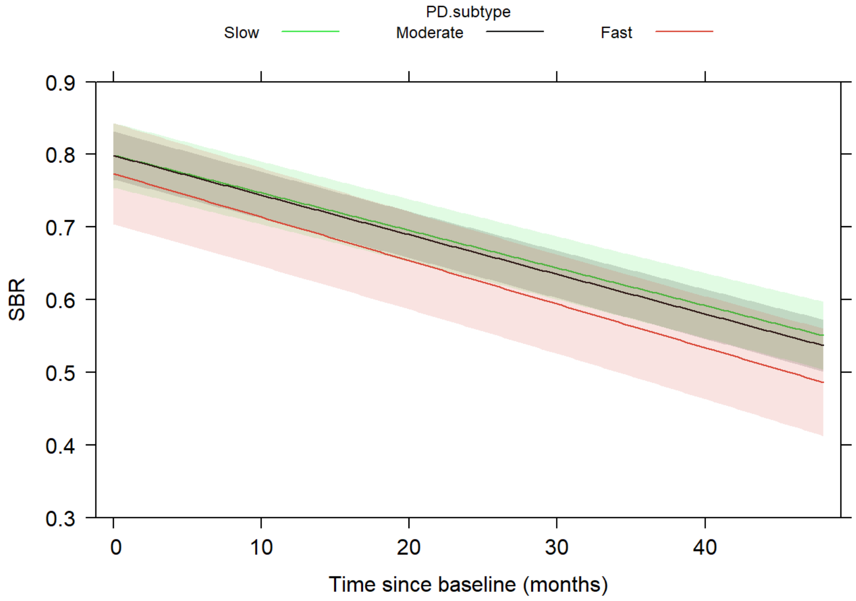


C. Whole striatum

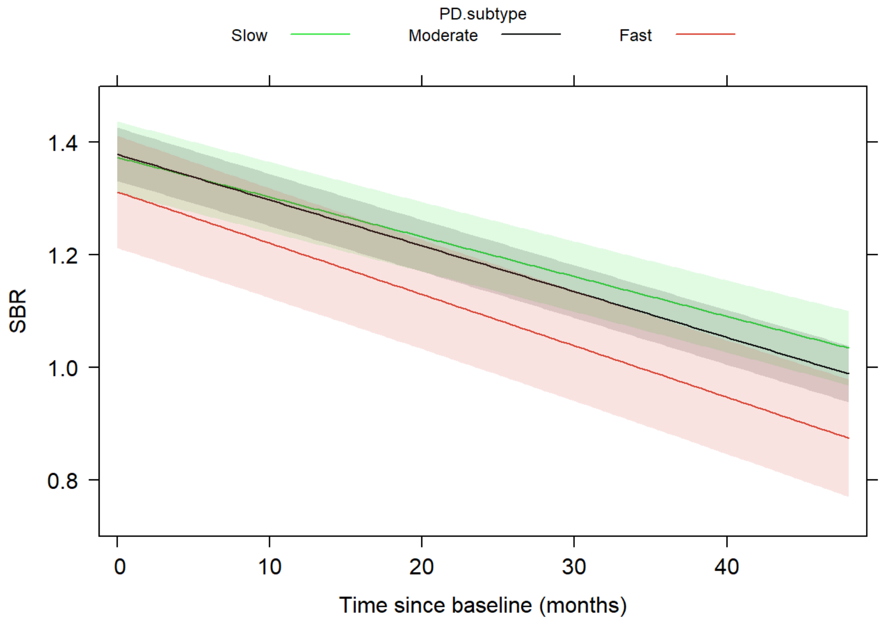


**Figure S5**: Longitudinal loss of dopaminergic neurons: Predicted effect plots of the modelled decrease in DaTSCAN specific-binding ratio (SBR) signal in the caudate (A), putamen (B), and whole striatum (C) using LMEM where time was fitted as a continuous variable and baseline signal as part of the response. The shaded area indicates the 95% pointwise confidence band for the fitted values, based on standard errors computed from the covariance matrix of the fitted regression coefficients.

A. Caudate

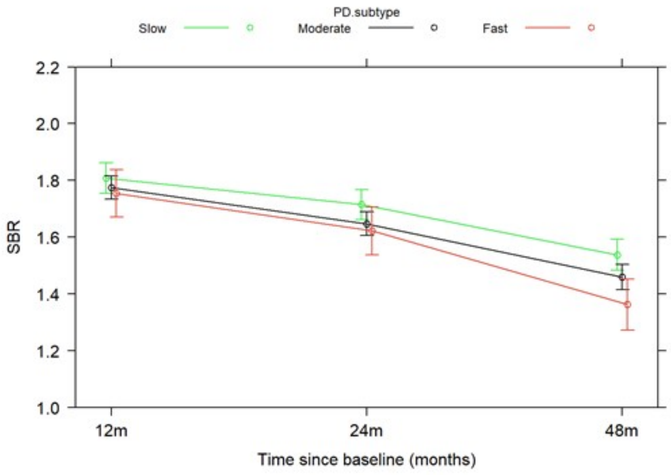


B. Putamen

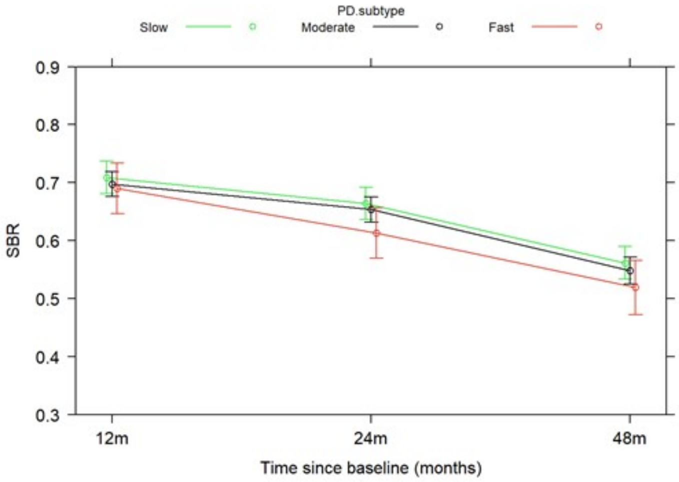


C. Whole Striatum

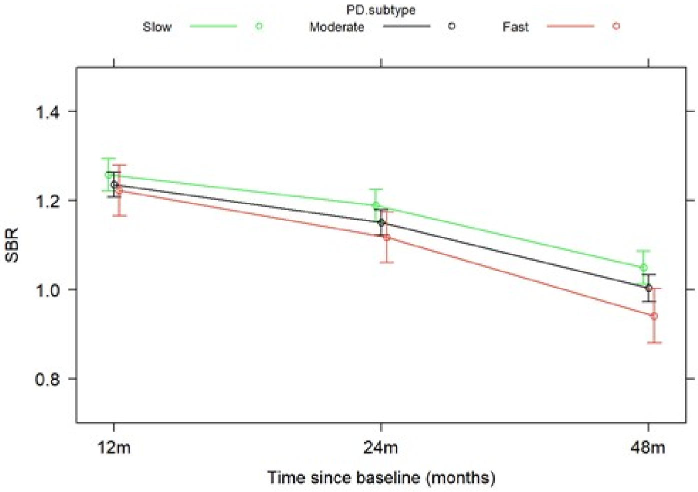


**Figure S6:** Longitudinal loss of dopaminergic neurons: Plot of predicted effects of the modelled decrease in DaTSCAN specific-binding ratio (SBR) signal in the caudate (A), putamen (B), and whole striatum (C) using LMEM where time was fitted as a categorical variable and baseline signal as covariate. The error bars represent the 95% confidence intervals, based on standard errors computed from the covariance matrix of the fitted regression coefficients.

### Machine learning reveals associations between variables and clusters

#### Sparse Group LASSO Penalised Binomial Regression

We applied a so-called intermediate data integration strategy [6] via a sparse group LASSO penalised binomial regression model (SGL) [7]. SGL is an extension of the classical LASSO originally introduced by Tibshirani [8]. SGL combines selection of individual features (like LASSO) with selection of entire feature groups, like group LASSO [9]. More specifically, the SGL optimization function has the following form:

$$\min_{\beta} \frac{1}{2} \left\| y - \sum_{l=1}^{m} X^{(l)} \beta^{(l)} \right\| + (1-\alpha)\lambda \sum_{l=1}^{m} \sqrt{\square} \,{}_{\square}^{\square}$$

where m denotes the number of feature groups (here: data modalities) and $\beta^{(l)}$ the coefficients for $p_l$ variables in group $l$. The first penalty (on group level) enforces a selection of entire data modalities. The second penalty (on feature level) in addition yields a sparse solution within selected data modalities. Hyperparameter α ∈ [0,1] controls the trade-off between both penalty terms. Hyperparameter λ controls the overall influence of the penalty. Optimal hyperparameter values were chosen via nested cross-validation.

#### Training Procedure

To train the models, the same patients were used as for the subtyping. The  extended set of 3472 SNPs was obtained via linkage disequilibrium based on 145 manually curated SNPs [10](see Section “Genetic variables and Pathway scores”). The full list of used variables and their grouping into data modalities can be found in the **Supplementary Spreadsheet**. The labels for training the models were the patients' assigned subtypes transformed into a binary setting (ie., one subtype being assigned 1 and the respective others with 0).

To develop a model based on our multimodal data, we considered several possible strategies (random forests, extreme gradient boosting, sparse group lasso), of which the sparse group lasso performed best and was further used for interpretation of predictor coefficients. All models were trained in a nested cross-validation setting with hyperparameter tuning and imputation occuring in the inner cross-validation. Imputation was done using the missForest approach [11].

#### Calculation of Mechanism Burden and Polygenic Risk Scores

For each mechanism $i$, we calculated a genetic burden score (stored in a vector $U_i$ of length N for all $N$ PD patients): We compute $U_i$ as $\sum_k M_{jk}$, where M is a matrix (dimension $N \times d_i$, length of $d_i$: number of all SNPs mappable to mechanism $i$) denoting the number of non-reference alleles of SNP $k$ in patient $j$. We then concatenate the $U_i$ to form a $N \times D$ matrix of $D$ mechanism burden scores for each patient. Additionally, we compute polygenic risk scores (PGS) $P_i$ for all PD associated PGS in the PGS catalogue [19] and PheWAS. For each PGS $i$, $P_i = M E_i$, where $E_i$ is a vector of effect sizes for SNPs from the PGS definition and M as described above. We then concatenate the $P_i$ to form a $N \times P$ matrix of $P$ polygenic risk scores for each patient.

### Cluster - Variable Associations Using Only Baseline Variables

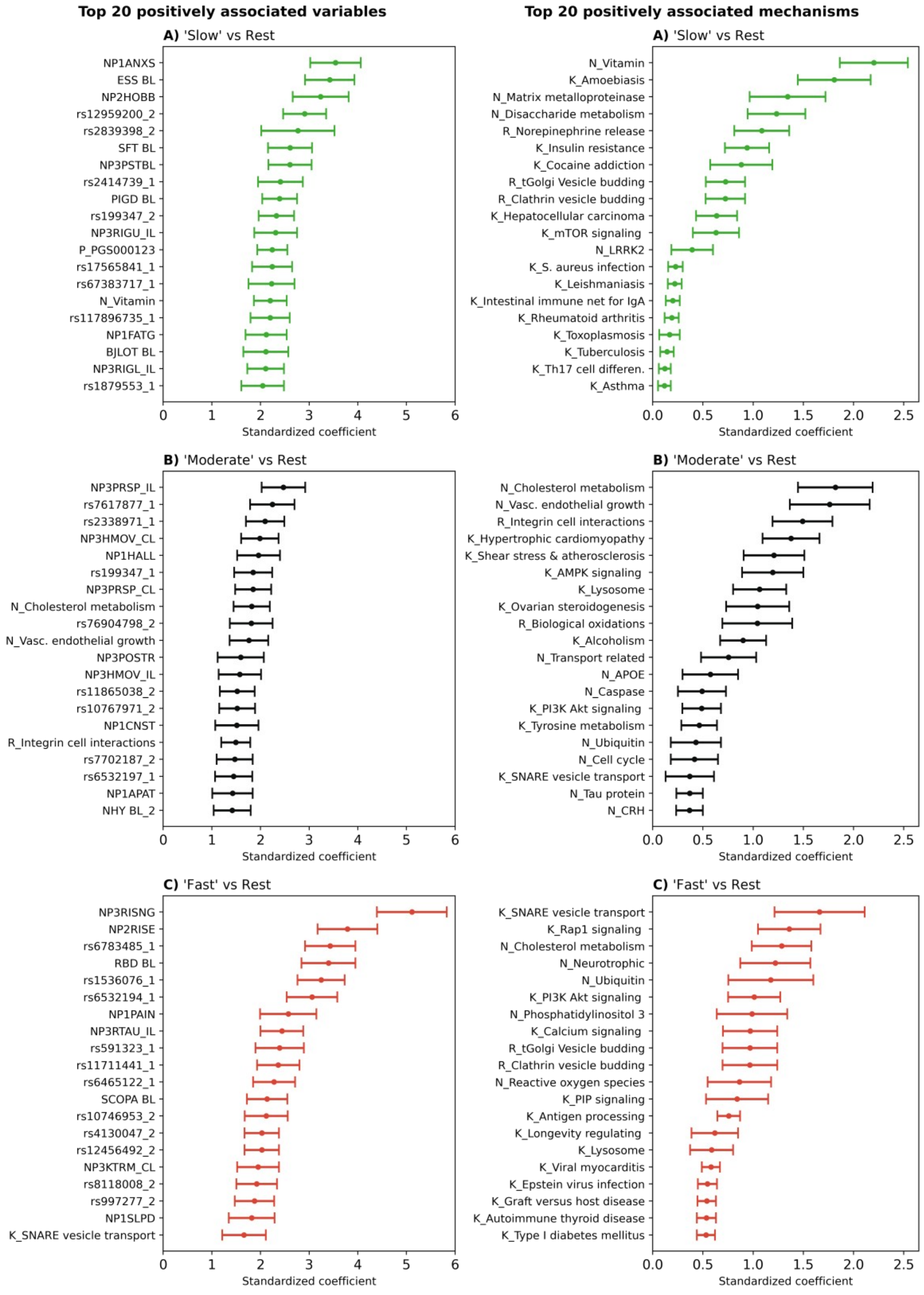


**Figure S7:** Top 20 variables predicting the respective progression subtype (model using only baseline data). The plots show the standardised coefficient for each variable. A stronger positive coefficient value in the plot indicates a higher likelihood of a patient to belong to the respective subtype. A-C, most predictive variables for ‘slow’, ‘moderate’ and ‘fast’ progression. Variables ending in ‘slope’ indicate the slope of the corresponding score measured after 3 months relative to baseline. The number after SNP IDs indicates the number of non-reference alleles. D-F, most predictive biological pathways. Pathways starting with ‘K_’, ‘R_’, or ‘N_’ originate from Kegg, Reactome, and NeuroMMSig, respectively. PGS denotes polygenic risk scores.

#### Differences in Response to Motor Symptom Therapy Across Clusters

##### Statistical Methodology

Separate linear mixed effect model for longitudinal data (LMEM) was fitted to the UPDRS 3 score data in the “ON”-state and in the ‘OFF’-state up to 60 months. The PD cluster was modelled as a categorical variable, time as a categorical variable, and baseline UPDRS 3 score data, as a covariate. To adjust for the dependency of the repeated observations within the individual, a subject-specific random intercept was fitted. The model was adjusted for age at baseline ( [30-55[; [55 - 65[; [65+[ years), disease duration at baseline ( [0-0.3[, [0.3-0.6[, [0.6+[ years), and LEDD at 9 months ( [0-300[, [300-1200[, [1200+[ ). No imputation was performed as data were analysed using a LMEM for longitudinal data, with embedded missing at random (MAR) assumption and implicit imputation. Diagnostic plots of LMEM models (fitted via R package *lmerTest* [20]) obtained via function residplot from the R package *predictmeans* can be found in **Figures S9, S10**. LMEM model was fitted using the R package *lmerTest*. Predictor effect plots were obtained using R package *effects*.

A. 'OFF'-state

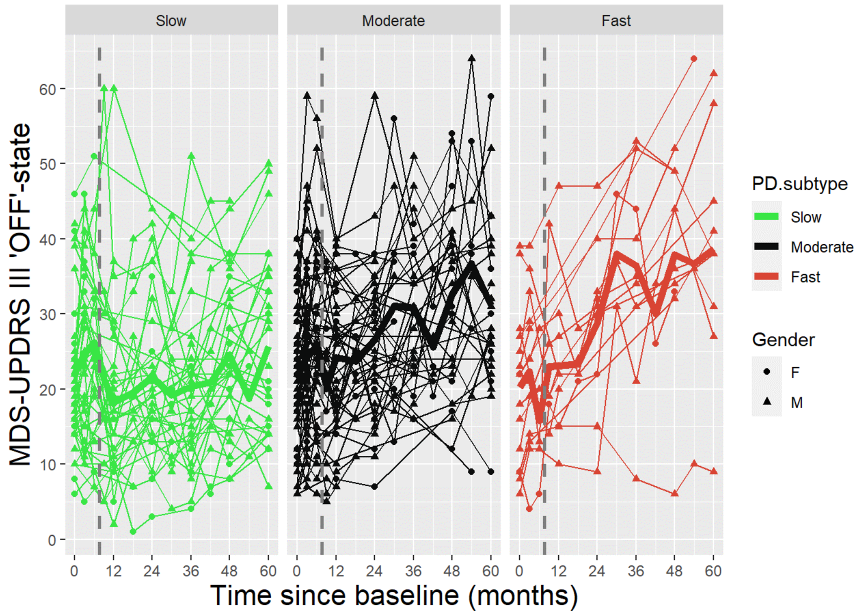


B. 'ON'-state

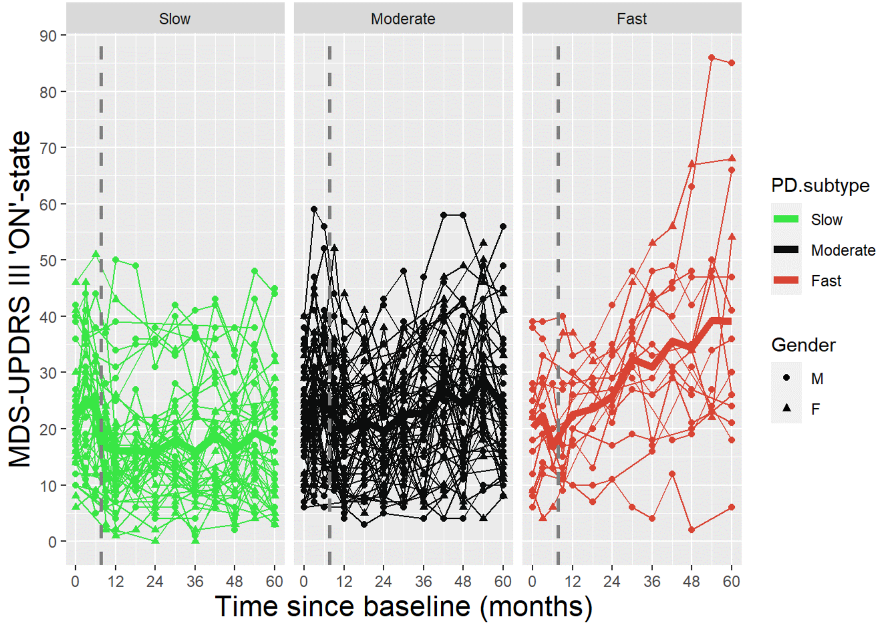


**Figure S8:** Individual line plots of UPDRS 3 score data in the (A) 'ON'-state and (B) 'OFF'-state among PPMI de Novo PD participants who initiated Levodopa or Dopaminergic agonist symptomatic treatment (ST) between 6 and 9 months post-baseline (N = 127) presented by PD subtype. M: Male (N = 88), F: Female (N = 39).

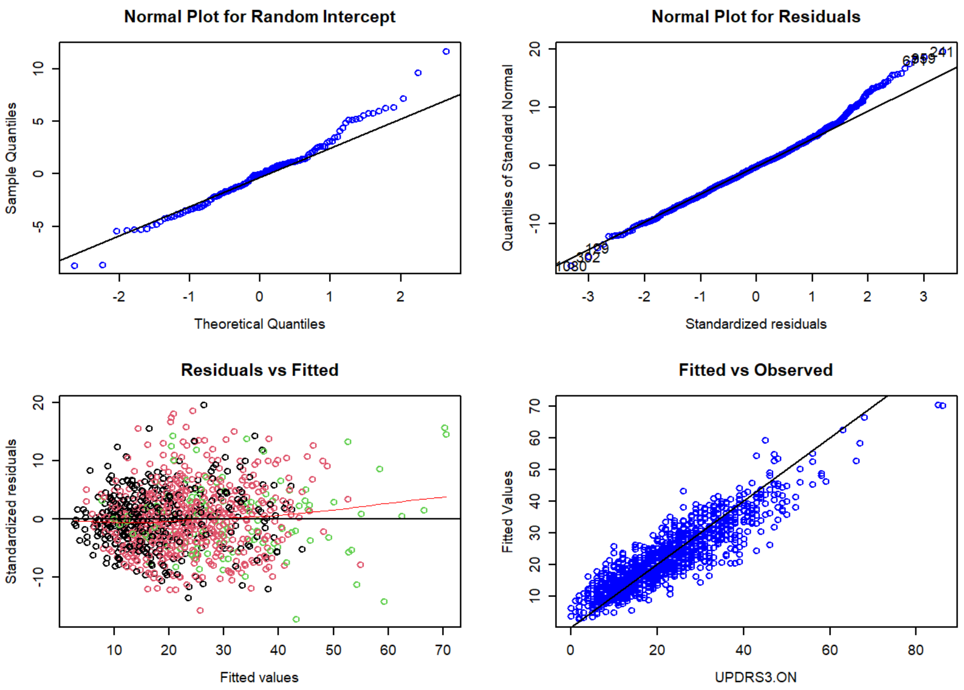


**Figure S9:** Diagnostic plots for the LMEM applied to UPDRS III **'OFF'-state** score data among de Novo PD PPMI participants who initiated symptomatic treatment (ST) between 6 and 9 months post-baseline (N = 121) where (A) time is fitted as a continuous variable and baseline score as part of the response in a piecewise model and (B) time is fitted as a categorical variable and baseline score as a covariate. For the 'Residuals vs Fitted' plot, datapoints are coloured according to the progressing group: 'slow' (green), 'intermediate' (black), and 'fast' (red).

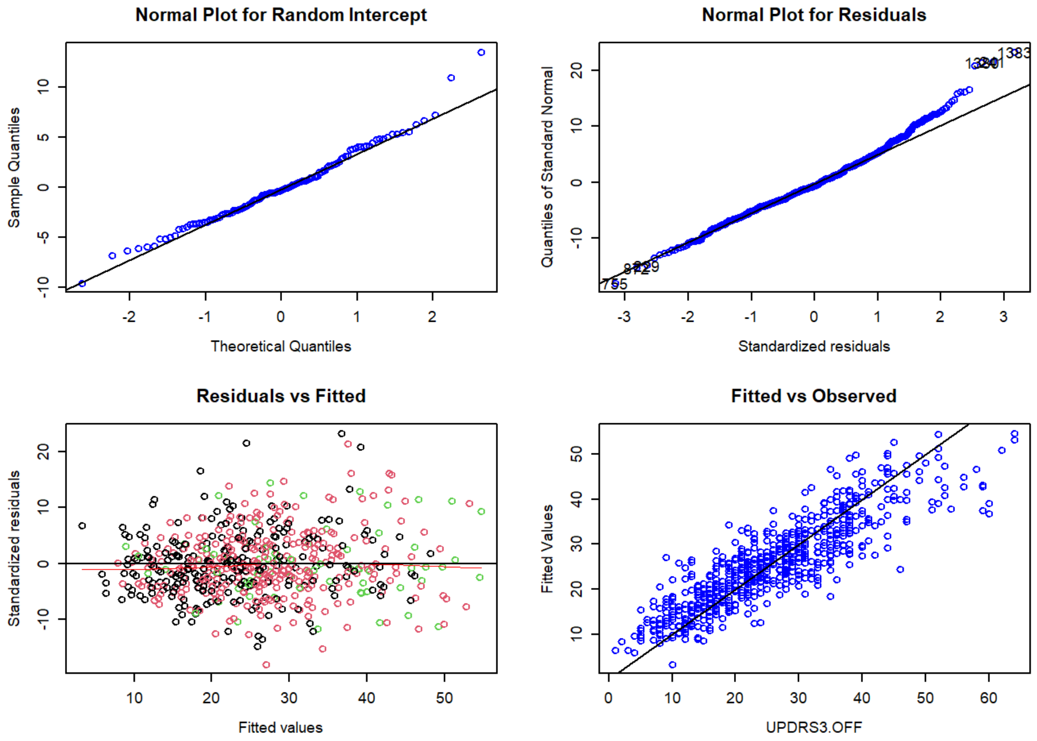


**Figure S10:** Diagnostic plots for the longitudinal Linear Mixed Effect Model (LMEM) applied to UPDRS 3 **'ON'-state** score data with time fitted as a categorical variable and baseline UPDRS 3 score data fitted as a covariate among de Novo PD PPMI participants who initiated Levodopa or Dopamine agonist ST between 6 and 9 months post-baseline (N = 127). For the ‘Residuals vs Fitted’ plot, datapoints are coloured according to PD subtype: ‘slow’ (green), ‘moderate’ (black), and ‘fast’ (red).

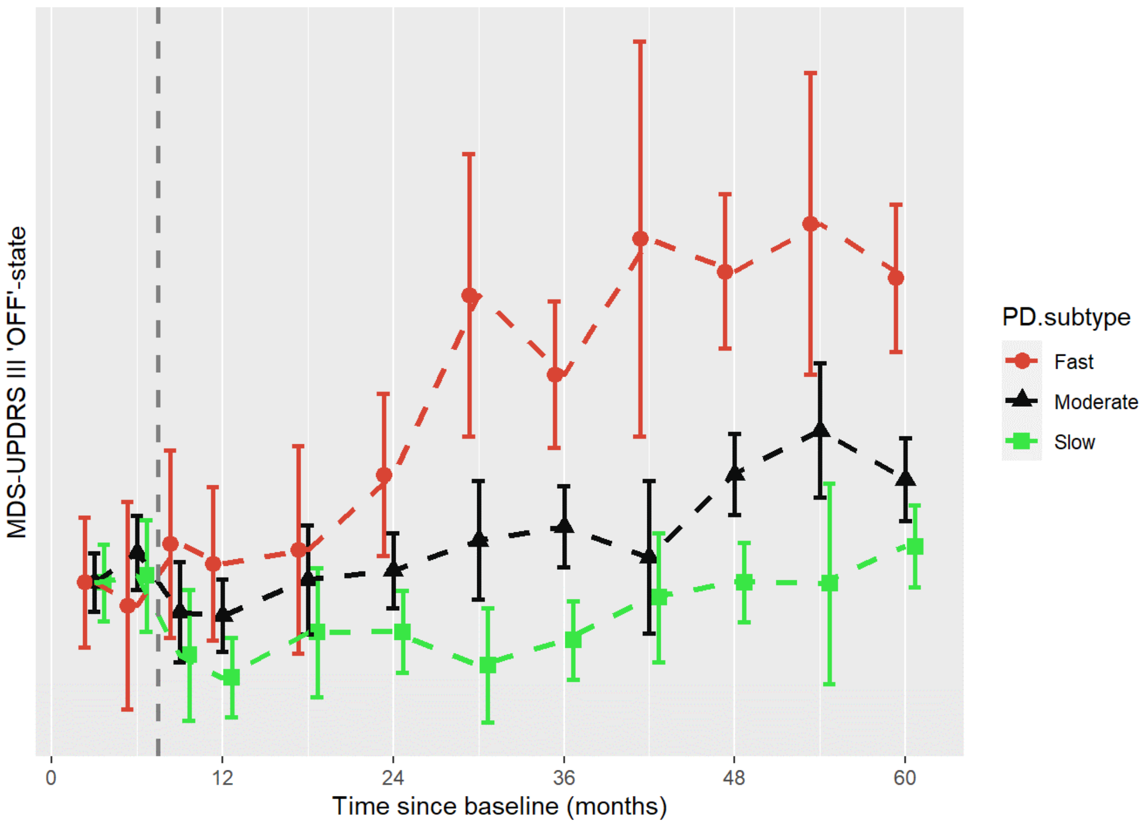


**Figure S11:** Differential response to symptomatic therapy: Effect plot of modelled UPDRS 3 ‘OFF’-state score progression prior to and after the initiation of Levodopa or Dopamine agonist in patients who initiated therapy between 6 and 9 months post-baseline using a longitudinal LMEM with time fitted as a categorical variable and baseline score fitted as a covariate. The error bars represent the 95% confidence intervals, based on standard errors computed from the covariance matrix of the fitted regression coefficients.

| ST type | PD subtype | % of PD subtype (count) |
|---|---|---|
| **LD** (+/- other) | Green / Slow | 38.1% (16) |
| | Black / Moderate | 53.1% (34) |
| | Red / Fast | 46.7% (7) |
| **DA** (+/- other) | Green / Slow | 57.1% (24) |
| | Black / Moderate | 40.6% (26) |
| | Red / Fast | 53.3% (8) |
| **LD and DA** (+/- other) | Green / Slow | 4.8% (2) |
| | Black / Moderate | 6.2% (4) |
| | Red / Fast | (0) |

**Table S1:** Type of ST prescribed among PPMI participants who initiated intake between 6 and 9 months post-baseline shown according to PD subtype (N = 127). LD: Levodopa, DA: Dopamine agonist.

| Visit | PD subtype | Count | LEDD (mg) | |
|---|---|---|---|---|
| **9 months** | | | Median | IQR |
| | **Green / Slow** | 44 | 225 | 115 |
| | **Black / Moderate** | 67 | 300 | 240 |
| | **Red / Fast** | 16 | 270 | 158 |
| **48 months** | **Green / Slow** | 44 | 503 | 400.0 |

| | | | | |
|---|---|---|---|---|
| | **Black / Moderate** | 67 | 400 | 253 |
| | **Red / Fast** | 16 | 478 | 415 |
| **60 months** | **Green / Slow** | 44 | 600 | 288 |
| | **Black / Moderate** | 67 | 590 | 296 |
| | **Red / Fast** | 16 | 710 | 453 |

**Table S2:** Levodopa equivalent daily dose (LEDD) in mg calculated as per PPMI protocol at the 9-, 48- and 60-month visits for de novo PD participants who initiated Levodopa or Dopamine agonists between 6 and 9 months after baseline (N = 127). IQR: Interquartile range.
